# Stakeholder Interviews to Inform Best Practice for Public Facing COVID-19 Wastewater Dashboards

**DOI:** 10.1101/2024.03.26.24304848

**Authors:** D. Morales, T. Rhodes, K.M. O’Reilly

## Abstract

**Background:** WW-based epidemiology is the detection of pathogens from wastewater, typically sewage systems. Its use gained popularity during the COVID-19 pandemic as a rapid and non-invasive way to assess infection prevalence in a population. Public facing dashboards for SARS-CoV-2 were developed in response to the discovery that RNA biomarkers were being shed in faeces before symptoms. However, there is not a standard template or guidance for countries to follow. The aim of this research is to reflect on how currently available dashboards evolved during the pandemic and identify suitable content and rationale from these experiences.

**Methods and Results:** Interviews were carried out with implementers and users of dashboards for SARS-CoV-2 WW data across Europe and North America. The interviews addressed commonalities and inconsistencies in displaying epidemiological data of SARS-CoV-2, clinical parameters of COVID-19, data on variants, and data transparency. The thematic analysis identified WW dashboard elements that can facilitate standardization, or at least interoperability. These elements emphasise communication among developers under the same organization, open access for identified stakeholders, and data summarized with a time-intensive graphic analysis through normalizing at least by population. Simultaneous communication of clinical surveillance is recommended. More research is needed on flow and faecal indicators for normalization of WW data, and on the analysis and representation of variants.

**Discussion:** WW dashboard development between 2020-2023 provided a ‘real-time’ iterative process of data representation, and several recommendations have been identified. Communication of data through dashboards has the potential to support early warning systems for infectious diseases.

## Introduction

WW-based epidemiology (WBE) is the detection of pathogens from wastewater, typically sewage systems. Indicators of presence/absence, or pathogen quantity are often measured using real-time qPCR, and provide a method for monitoring infection in a population. The basic mechanism behind this is that many infections replicate in the gut and so genetic material can be detected from environmental material with faecal contamination, especially sewers. While WW data and analysis continues to be a developing field of public health, it has been used to detect pathogens since at least the 1930s.^1^ The best known ‘use-case’ is in polio eradication, where poliovirus can be detected in sewage from infected communities, and if local transmission is thought probable timely vaccination responses can prevent poliomyelitis cases, such as that reported in Israel in 2013.^2^

Today, WW is being applied to detect levels of illicit drug use, pharmaceutical consumption, antimicrobial resistant microbes, chemical exposures, and infectious diseases.^3,4,5^ WW data can provide additional critical information that can be used in conjunction with other methods of surveillance, such as clinical surveillance. A concern with exclusively relying on clinical records for surveillance is that the data collected from health centres may be underreported and biased, and will not detect asymptomatic infections.^6^ A promise afforded by WBE is that when reported and analysed with clinical data, researchers are provided with health information that *fills in the gaps* regarding how a virus moves through the population while also analysing variants.

In April of 2020 it was found that SARS-CoV-2 (the causative agent of COVID-19) could be detected in the stool of active COVID-19 cases. In fact, RNA biomarkers of SARS-CoV-2 were being shed in faeces days before COVID-19 symptoms developed. Researchers in the Netherlands reported the first example of WW surveillance that same April.^7^ Several studies have illustrated a positive correlation of viral concentration in WW with the number of SARS-CoV-2 cases, and many settings have established WBE to approximate the disease burden of COVID-19 in communities.^8–10^ This development has been especially useful while health systems were overwhelmed, and documented examples of public health actions based on WW data are now emerging.^11–13^

During the COVID-19 pandemic there was a demand for countries to rapidly develop and communicate surveillance data via online dashboards.^14^ Dashboards are a public facing display, typically an online webpage, that provides a data summary for stakeholders. While many disease-specific^15,16^ and general dashboards^17^ existed prior to the COVID-19 pandemic, engagement with dashboards was typically limited to subject experts. That COVID-19 was an epidemic which affected everyone and was a rapidly evolving situation, meant that there was a demand for real-time information that would indicate community spread and contextualise mitigating actions.^18^ With reference to dashboards designed specifically for WW data, dashboards were developed without a unifying organizing body and there was no template, leading to variability in reported results, data standards, meta data requirements, and quality assurance. An analysis of dashboards present on the COVIDPoops19 site emphasised the distinctions: in 2022, 49% of the dashboards represented data in the form of graphs while 48% presented maps.^19^ How data is represented, and variabilities in data visualisations, may shape how evidence comes to be known and understood among different stakeholders, both among the scientific community, policy makers and community members. The public communication of science in the COVID-19 pandemic was at times characterised by multiple and contested interpretations of visually represented data, emphasising the complexities of translating data in early warning efforts and emergencies.^20^ The swift implementation of WW surveillance may have caused uncertainties in the reliability of the data. As argued by Rhodes and Lancaster,^21^ the methods of presenting data may be as important as the data itself and can thus be considered as a form of evidence making. Consequently, an understanding of how dashboards were developed is an important part of public health.

The aim of this research is to reflect on how currently available dashboards evolved during the pandemic to understand stakeholder and implementer perspectives on the preferred content and rationale for SARS-CoV-2 WW dashboards. Further, we are interested specifically in whether such preferences align in ways which enable the standardization of SARS-CoV-2 WW dashboards.

## Methods

Research on a sample of online SARS-CoV-2 WW dashboards was conducted to develop research themes and interview questions. Both the EU Sewage Sentinel System for European Dashboards^22^ and COVIDPoops19^23^ were used to identify specific WW dashboards. These websites were put together by research stakeholders to act as a hub for stakeholders view independent dashboards. Our initial review of dashboards focused on the granularity and presentation of WW data and the inclusion of clinical data. This gave rise to questioning how and what epidemiological data should be hierarchized in dashboards. We therefore proposed a qualitative interview study of stakeholder perspectives on this question.

Participants were recruited from organizations with known dashboards, through informal networks, and via social media. We adopted a purposive sampling approach in order to include a variety of backgrounds and forms of expertise, including among researchers, health providers, public health workers, government employees, and academics across Europe and North America. There were no prior relationships with the study participants and the interviewer.

Participants were asked to fill out a consent form, including questions on demographics and basic information to help with interviews. Demographic questions included gender, country of work, country where WW data are collected. To help with interviews, we requested a link to the dashboard used in their country of work, their profession, how long they have worked in this role, and if they identified as a Developer/Implementer of dashboards or as a User/Stakeholder. The interviews were conducted over Zoom and were transcribed by a transcribing software. In preparation, a pilot interview was carried out, in which it was identified that a PowerPoint presentation would be useful to guide the interviewees through the questions.

Interviews consisted of a 30–45-minute semi-structured interview, carried out throughout August 2023 by DM, who was carrying out this study as part of her MSc, and this was explained to all study participants. Only the interviewer and interviewee were present at the call. The interview guide was separated into three sections (Appendix 2):

1. Participants were asked about the experience and process of creating the dashboard. The question was designed to explore how often dashboards were accessed and analyse the strengths/weaknesses of current design.
2. Participants were asked how epidemiological information should be hierarchized for the purpose of standardization. The second part of this question focused on what WW units are preferred to represent RNA copies of SARS-CoV-2.
3. Participants were asked about the content that should be included alongside WW data and the transparency of this data. Participants were asked for insights on clinical data and variant surveillance, how accessible dashboards should be and for whom.

Following the transcription of the audio, the transcriptions were uploaded into NVivo, a qualitative coding software. Codes were developed based on the Braun and Clarke Thematic Analysis.^24^ More specifically, a theoretical thematic approach was used to code sections that were relevant to the original objectives, and an open coding process was used to allow for modification. Transcripts were not returned to the interviewee for comment.

The original parent codes included: experience with dashboards, process of development, gaps of knowledge regarding the content within a dashboard, epidemiological hierarchization of the graphic, and epidemiological hierarchization of WW units of measurement. Within these parent codes, sub sections were created based on the frequency and importance of a section.

Participation in the survey was voluntary, with informed consent, and all analyses were performed on anonymized data. This study was approved by the ethics committee of the London School of Hygiene and Tropical Medicine (Ref: 28778).

## Results

### Demographics of Study Participants

A total of 32 people were contacted via email, with 14 interviews conducted, resulting in a response rate of 43.8%. Reasons for non-participation were non-response or not being able to participate (interviews took place in August, a vacation period). No repeat interviews were required. Participants were based across Europe and North America, and worked in 10 countries, but collected WW data from 15 countries, including within Africa and Asia. Seven participants identified as both a User/Stakeholder and Developer/Implementer while five identified as just a Developer/Implementer and 2 as just a User/Stakeholder (Table 1). Participants had worked with WW data for an average of 12 years.

**Table 1:** Demographics of Study Participants.

| Country of Work | Participant<br>s | Interaction with WW dashboards |  |  | Mean<br>years<br>worked<br>with WW<br>data |
| --- | --- | --- | --- | --- | --- |
|  |  | User/<br>Stakeholder | Developer/<br>Implementer | Both |  |
| **Austria** | 2 | 0 | 0 | 2 | 2 |
| **Finland** | 1 | 0 | 1 | 0 | 20 |
| **Ireland** | 2 | 0 | 1 | 1 | 14 |
| **Italy** | 1 | 0 | 0 | 1 | 25 |
| **Netherlands** | 1 | 1 | 0 | 0 | 21 |
| **Norway** | 1 | 0 | 1 | 0 | 3 |
| **United Kingdom**<br>**(Northern Ireland,**<br>**Scotland, and Wales)** | 4 | 1 | 1 | 2 | 7 |
| **United States of**<br>**America** | 2 | 0 | 1 | 1 | 3 |
| **Total** | **14** | **2** | **5** | **7** | **12** |

**Table 2:**
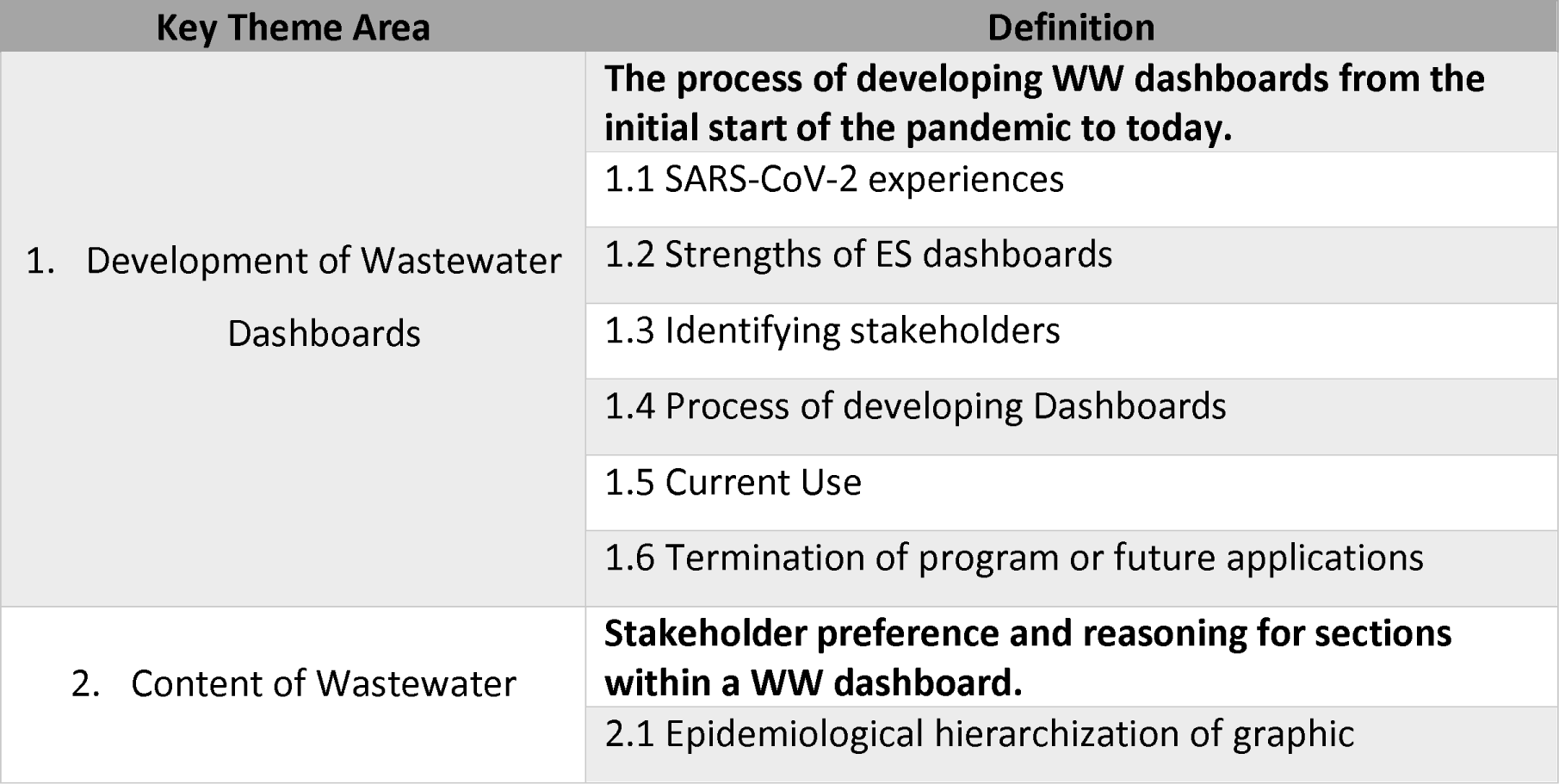
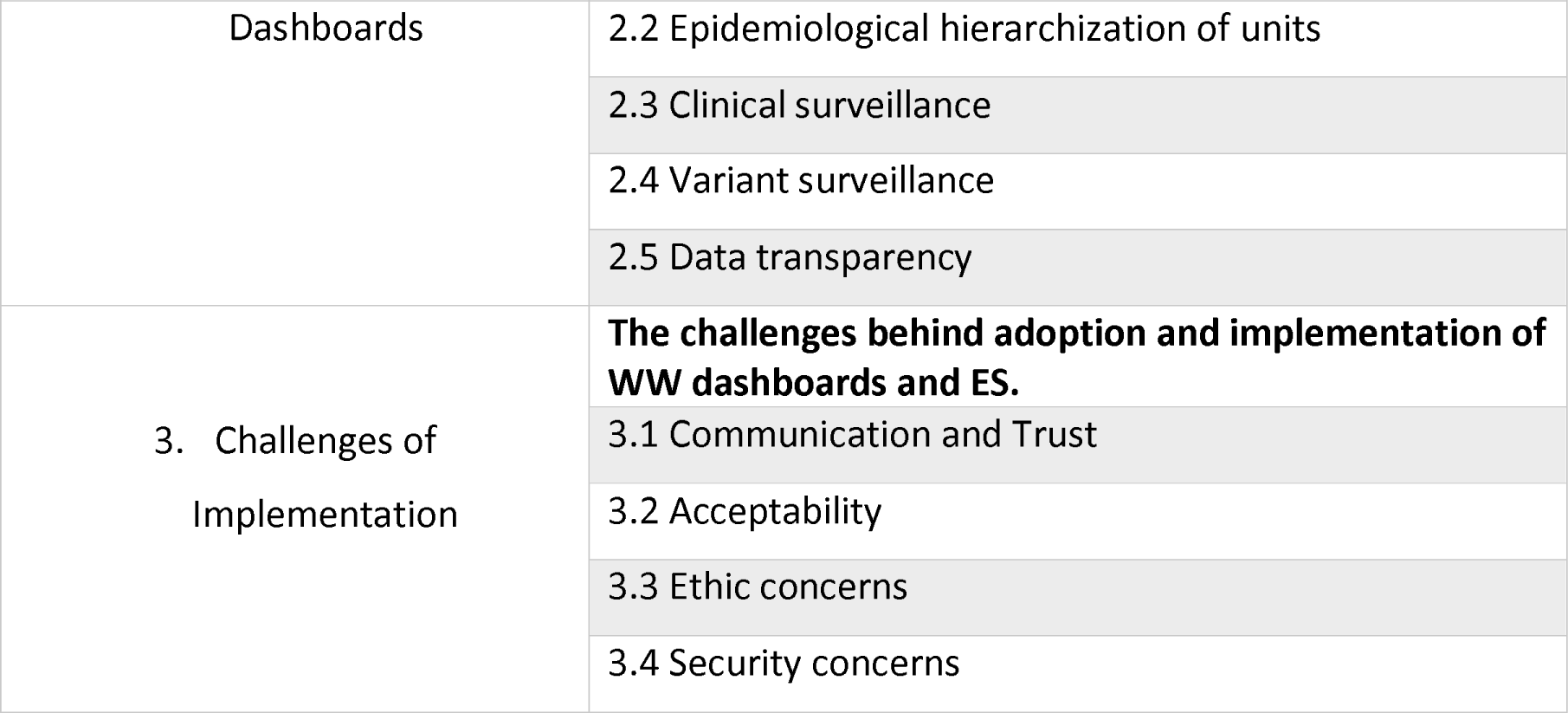
Themes from the Braun and Clarke Thematic Analysis coding process.

There were three identified themes from the coding process: the development of WW dashboards, the content of WW dashboards, and the challenges of implementation.

### Theme 1: Development of WW Dashboards

#### 1.1 SARS-CoV-2 Experiences

While some countries did not begin investing in WW surveillance until the COVID-19 pandemic many others already had WW surveillance in place and were able to transition their equipment from a previous different target to SARS-CoV-2. Only four participants mentioned WW surveillance that were already conducted in their countries, with applications including polio, illicit drug use, faecal contamination of bathing waters, and specific bacterial/viral projects conducted through academic institutions. Eight participants did not start working with projects focused on WW surveillance until 2020. One participant mentioned that while dashboards had existed pre-COVID-19, *“…the technical reason for why we didn’t develop more, was that we didn’t have a good reason for deploying them*”. In fact, when the Scottish government decided to conduct routine monitoring of the SARS-CoV-2 pathogen in June of 2020 this was the first national surveillance system for a pathogen that had ever been done independently of the UK.

During the initial days of the pandemic, WW surveillance systems were put together very quickly. A developer explained that,

> “*The WW surveillance was novel because while it did exist before for polio or other applications, it was novel in its bipartisan approach because it brought in research and academia*”. (3)

One participant described how their team discussed and promptly executed initial research without securing funding. These unprecedented circumstances were new territory for countries and a stakeholder described it as if “*Everybody was trying to invent the wheel*”, and many others expressed that there were no standardization or protocols for countries to follow. Nevertheless, experts were communicating between countries, and the European Commission supported member states. Two participants working in government appointed health roles shared that until the intervention of the EU many governments were not interested in WW epidemiology.

The first step that developers took when designing dashboards was to identify the audience and stakeholders. For example, an epidemiologist might prefer exportable data to create models themselves, while government/health policy makers require a plain explanation of the data. All participants reported that the dashboards were not originally designed for the general audience, and instead for professionals such as national health security and safety agencies, local health authorities, researchers, and policy makers.

All participants expressed the common theme of dashboard creation following an iterative process. As one developer stated, “*The dashboard had to be correct in all regards so to reduce the corruption, or the possibility for corruption*”. Each developer explained that the ideal process was done in stages with user feedback present at each stage, and four developers credited team collaboration for success. Only one developer mentioned the use of specific guidelines that were followed, and this was the Government Design Principles.^25^ Two developers mentioned using the ONS coronavirus survey carried out in the UK as an exemplar for data communication.^26^ One user expressed that their dashboard could not succeed until statistical support was sought out for analysis of the data. Two developers stated that a major improvement was simply having more data points, one of them stated, *“…we first only had qualitative data detected and not detected, and later on we were able to solve for trends and quantitative results*”.

Challenges were encountered during dashboard development, such as balancing data privacy and clarity, “*What should be the resolution? So the provinces that was in the end allowed, but not the communities*.” The presentation of data evolved during the pandemic, alongside there being more data to display. Several participants commented that explanations of the data, and guides for users of the dashboards were included in the later stages,

> “*There are more information now than at the beginning, more data points, more information in general [and] more explanations. What the data means and how to use the data*” (1)

As users were engaging with the data, use varied from checking the dashboards weekly, to not at all. Three users explained that they rely more on weekly reports provided to health officials instead of checking the physical dashboard. A stakeholder explained that the Center for Disease Control (USA) dashboard provides automated updates for users so even if the dashboard is not checked daily, it will still contact stakeholders. Provision of dashboards to a wide audience also presented research opportunities,

> “…*there are so many interesting people who are starting to ask questions, and these questions are helping us to see and consider different things. Having this feedback helps a lot, especially in the beginning. When we were developing, no one had done it prior*” (8)

As of August 2023, many countries have either started to switch off their dashboards, terminate programs, or have taken a hiatus in the use of WBE. In fact, a couple of weeks before the interviews, the Welsh government discontinued the development of the dashboard and stopped clinical surveillance of SARS-CoV-2 (although some moderate data collection has since resumed). The discontinuation of SARS-CoV-2 WW data collection has encouraged researchers to think beyond COVID-19, for example by expanding towards a multi pathogen surveillance system including seasonal respiratory and gastroenteritis diseases,^27,28^ as well as interest in monitoring for antimicrobial resistance.

### Theme 2: Content of WW Dashboards

#### 2.1 Presentation of WW data and associated metrics

When asked about how users would hierarchize epidemiological data in terms of a graphic, eight stakeholders explained that the dashboard of the country that they worked in used a virus particle-time line graph representation. These same stakeholders considered this the most valuable information to include. The most popular reason for this preference was that they valued seeing how the virus load changed over time. One user stated that,

> “*I’d say most users derived most benefit from looking at it in a line graph and looking at the fluctuations in the data…It was easier to tell a story with the line graph*”. (2)

The only challenge described was that sometimes line graph representations can be over too much time. One user expressed that,

> “*The problem of this graph for me at least, is that it is for a very long time, so most often the stakeholders are interested in the last couple of weeks*”. (4)

Only two participants preferred the use of a geographical map. The reason for the geographic preference was that these stakeholders preferred to look at the bigger picture, a stakeholder expressed that, “*… we have an eagle’s eye view. We look from the top, so, we are interested in the occurrence of the presence of the virus*”. Although it was common for participants to compliment the design of the map, they admitted that it was not realistic globally, a stakeholder explained that, “*It depends on the infrastructure that is there, in the European regions, there’s nice coverage of networks, but that is not the case in all locations of the world*”. For many countries the information that needs to be included in a geographical display is either not public data or not part of a public system. In fact, many challenges were expressed about the design of a map and the most common being that the data changed too much depending on population level leading to changing boundaries. It is very difficult to show this data in one graphic especially in large countries, a developer explained that,

> “*Our problem with the map is that in […] we have a huge diversity in the size of our treatment plants. So, they range from serving from 1000 people up to 5 million and when you have that kind of diversity these numbers are no longer comparable*”. (9)

When stakeholders were asked about measuring the units of SARS-CoV-2 in WW, even with varying levels of expertise, every stakeholder emphasized the importance of normalizing the data (ie. at least accounting for population size in the units of measurement reported). Thus, each of the stakeholders ranked the SARS-CoV-2 gc/L as the lowest priority, and one developer stated that, “*I wouldn’t be very comfortable showing non normalized data at all, because of misinterpretation*”. Five stakeholders identified normalizing by population as their top priority, and in fact it was the most common unit used throughout each of the dashboards. However, it was identified as a challenge when the population is not mapped against a sewer shed and it might not work with diverse or temporally changing population sizes. After normalizing by population, stakeholders preferred normalizing by WW flow. This was identified as beneficial because it corrects for rainfall and other contaminants in sewer systems. Next, it was preferred to normalize by a faecal indicator. However, there were mixed responses on the reliability of this method. While three stakeholders prioritized this unit in their dashboards, others countered the benefits and one stated that,

> “*There are several options for these [faecal] indicators, and the current challenge is that there is no global gold standard. So that it seems that any indicator might be good, but there are different options that different groups are using and that’s the main challenge at this point*.” (4)

another stakeholder said,

> “*We know that correction improves the statistical scattering, and it compensates, but none of the current faecal indicators are really capturing the picture as it should, presumably because you may need a whole range of faecal indicators to cover it*”. (3)

Nevertheless, the most common faecal indicators mentioned were the pepper mild mottle virus (PMMOV), ammonia, and crAssphage. Lastly, two stakeholders valued the importance of the qualitative metric. One of these stakeholders expressed why they chose this method by explaining,

> “*It is robust enough to be interpretable overtime at different states, whether concentrations are high or low. And it’s interpretable across our jurisdictions, so that’s what we use for our sort of initial like just how the jurisdiction is doing? qualitative is the answer*”. (9)

One developer explained that their dashboard compares by percent change metrics instead to discourage comparisons among sites.

#### 2.3 Clinical Surveillance

The question of including clinical surveillance in dashboards was unanimously described as valuable. In fact, the only reason that it was not included in certain dashboards was due to a lack in funding. Most stakeholders claimed that clinical surveillance aided in validating the WW data and would prefer to be able to plot the case data alongside. In fact, one participant emphasized the fact that,

> “*WW based surveillance is always additional information. Don’t use it stand alone. I mean, even if you have a dashboard, it doesn’t mean it is the solution to all your problems…the first rule in […], it is always supplementary*”. (3)

Nevertheless, it was frequently stated that under-reporting from clinical surveillance has increased in recent months, this has resulted in WW data has becoming the early warning system it was originally proposed to be, and in some circumstances the sole source of information.

#### 2.4 Variant surveillance

When users were asked about their opinion on variant surveillance, users unanimously mentioned the importance of conducting the research, however, they were divided on including this information on a dashboard. The answers were dependent on identifying the audience of the dashboards. Nine users prioritized the importance of it, while five expressed concerns with including this type of data. Many users shared that they thought variant data was more academic than informative, others claimed that genomic sequencing is not advanced enough to validate the data, and others said that they are not sure about including this data. One developer explained their hesitancy by explaining, “*You’re not treated any differently based on what variant you have. Your doctor’s making the same decisions. We’re recommending the same protective actions*.” Regardless of these reservations, every stakeholder in support of including variant surveillance emphasized the importance of communicating the information to the users in an accessible language to avoid confusion.

#### 2.5 Data Transparency

The topic of open dashboards versus restricted dashboards had varied opinions among participants. Overall, stakeholders emphasized the importance of having open data transparency for everyone. It was frequently mentioned across interviews that if taxpayers are paying for the program, then they should have access to the data. Although no stakeholder explicitly mentioned restricting data, many stakeholders described having to be cautious with sharing data openly. Some stakeholders cited government hesitancy and confidentially concerns and explained that they have restrictions with sample size. For example, a stakeholder explained,

> “*We have a certain rule that there needs to be enough people in these WW sample locations. Usually, it is more than 20,000 persons before we publish the data. Also, if the case numbers are lower than five cases per community, we are not publishing those small number of cases*”. (4)

Similarly, in the European dashboard the site is restricted because of data agreements with member states. In this case the data provider remains the owner of the data. One stakeholder explained that the raw data is restricted because “*You don’t want someone taking your data and doing open manuscript and publishing before you actually get the chance to do it*”. Two stakeholders mentioned having to separate the data into an internal dashboard and a public dashboard. In fact, a developer explained this separation by stating, “*One of the big failings initially was we tried to make a website that would work for everyone, and it worked for no one*”. Regardless of the current dashboard status, most of these data transparency perspectives can be summarized by the following comparison stated by a stakeholder,

> “*In case of SpaceX, they give the public the information they need to understand what’s going on, and then there’s a lot of technical data underneath and unless you’re an expert, it doesn’t really inform you at all. Or even worse, it might muddle the waters*”. (7)

### Theme 3: Challenges of the Implementation of Dashboards

#### 3.1 Communication and Trust

The main challenge identified across every interview was minimizing the risk of misinterpretation. Developers needed to report the data in a language that everyone who had access to the dashboard could understand, and a developer stated that,

> “*Not everybody’s going to have expertise in WW data interpretation. So even when sharing it within public health it needs to be clear, and even when communicating to our senior management, we need to make everything very clear as to what the signals are saying, and what we can’t say from the signals*”. (5)

One stakeholder explained that dashboards have global interest however, for a long time their dashboard was only in the national language and translations were included to meet the demand of international interest. Another communication challenge was updating dashboards. Dashboards need to explain a potential lag in data reporting or other delays, and one developer stated that “*We’ll occasionally get questions from health departments about why our internal dashboard shows one thing, while the COVID Data tracker is showing something different*”.

The main concern with communicating data based on geographical locations is that while It’s great to have a non-specific catchment, if that catchment falls over multiple different health boards or different geographies of interest, the information is diluted which limits interpretation. The representativeness of a site for a specific locality and the coverage achieved was difficult to communicate for many audiences. A developer stated that,

> “*People will look at this measure and say in my big, 4 million town, my WW is currently measuring 100,000 particles per 100,000 people. And in this tiny little town, it’s also 100,000 per 100,000 people, what are they doing wrong?*”. (9)

Interest from clinicians or in public health specialists was challenging in some settings. A developer explained this hesitancy by stating that “*This happens every time you have something new and innovative…you have to prove the value*”.

#### 3.2 Acceptability

Initial development of WBE perhaps lacked engagement with people working in public health. One stakeholder stated that, “*The link or collaboration with public health was missing and very few of the people at the monthly meetings were from public health backgrounds*”. Instead, many of the original stakeholders were people with environmental backgrounds and many participants claimed that these two worlds were not communicating effectively. A common theme identified was that WBE only works if the health sector is the implementor. This theory was exemplified by a stakeholder stating that, “*The WW people, they are very helpful, but they are scared that they have to do the job, so in other words that they would have to pay…*”.

In the case of acceptability many stakeholders explained that there was discourse between clinical and WW teams, and one implementor stated “*There is also a lobby of the medical sector, who want to maintain their unique position in society for doing disease surveillance*”. Overall, stakeholders mentioned that initially the health sector was not willing to accept this different type of surveillance. A stakeholder explained their frustration by claiming that, “*There are too few doctors anyways and they don’t have time, and they don’t need to have this bureaucratic burden…This additional burden*”.

During the pandemic many practitioners were not utilizing dashboards because they did not understand what dashboards added to their practice. A stakeholder explained that the practitioner’s perspective was that,

> “*I already know there’s COVID here and I already know what’s going on, how is knowing that going to help? and that’s not untrue, for someone in a hospital, I don’t know that they need to include WW as part of their thinking and diagnosis*”. (9)

Although, that perspective was the case for COVID it was stated that as WW surveillance moves to other pathogens the perspective of WBE and dashboards may change because of the potential to inform clinical care.

#### 3.3 Ethical and Security Concerns

A common theme across the interviews was that with WW sequencing individual cases should not be identifiable, a developer stated that,

> “*This information can be abused very easily, so the decision of what goes to the public and how it goes to the public, and how you’re communicating it is a very sensitive one, you have to protect vulnerable groups*”. (3)

Privacy concerns were stated to be more of an issue with small catchment areas such as with university monitoring.

In the United States, since 9/11 WW systems have been identified as a potential target of a biothreat. As a result, geolocated data is considered a privacy concern and thus WW information is not public data. Another stakeholder specified that there are legal implications behind releasing WW data, for instance “*Who owns that WW and consequently the information encoded?*” The conclusions that can be drawn from WW are advanced enough to be able to say something about health-related risks, and thus stakeholders are worried that interest from the private sector could affect life insurance or healthcare insurance.

## Discussion

WW dashboards for SARS-CoV-2 became an integral part of communicating information during the COVID-19 pandemic, and as individual testing reduces in frequency, WW dashboards have become the only regularly updated information on COVID-19 in many countries. To our knowledge this is the first attempt to interview developers and stakeholders of WW dashboards for COVID-19. We identified that the use of WW dashboards were in many cases unprecedented and so much was being learnt during the development process. The intended audience for dashboards were stakeholders involved in public health across a wide spectrum, where viewing time trends was the most useful output. While dashboards were not initially intended for a general audience, it became apparent that the public were broadly interested in the information displayed. A recognised challenge is identifying the most appropriate units of virus concentration for WW data, as there is currently no consensus on the minimal data required. Appropriate methods for flow normalisation are needed to improve interpretation of time trends and to support comparison across multiple sites and locations.

### Innovation and experimentation

The WW surveillance experience that countries had before the start of the SARS-CoV-2 pandemic impacted not only how quickly dashboards were developed but also the reliability and acceptability by policy makers. We found that until governmental bodies were convinced that WW surveillance could be a valuable method many academic institutions were operating with their own resources. Even if funding was provided, researchers were operating on temporary support because the duration of the pandemic was unknown. The methods of data collection and the metrics included within these dashboards were fully dependent on the resources available to these teams. The lack of resources may have contributed to methodology gaps in sampling and analysis of the WW. Looking forward, we note that continued funding for infectious disease surveillance using WW varies across settings, with considerable investments in the USA^29^ and the European Union,^22^ and an uncertain funding environment from charitable and governmental research funding bodies. Both the collection and presentation of WW surveillance data are important to improve and refine, both for endemic diseases and in advance of the next pandemic.

### Data presentation

The data displayed on a dashboard is dependent on the question that stakeholders are seeking to answer, which may be stakeholder dependent. However, there was a strong preference to observe trends in time along with comparisons within a local and/or national area. The second preference for display is through a geographical map. This graphic is dependent on the availability of catchment area data, how willing a country is to show sewer-catchment sites and the limits/boundaries of these systems. This information is crucial for policy makers that are interested in comparing the presence and intensity of the virus to other regions. There is a tension in the detail of information provided: on the one hand providing dashboards with up-to-date granular detail of infection trends has been described as a form of democratisation where failures in the social safety net can be seen and addressed,^30^ and in this study the open data approach facilitated innovation, and on the other hand this granular detail may be seen by some as a security risk and could have implications for healthcare access. Community, stakeholder and policy engagement is required to decide if the democratisation of data outweighs the perceived risks of making data available.

Even though stakeholders prioritize normalization of units, the specific factor of normalization is dependent on the data collected, and the technology available. Normalizing by population recommended to compare across sites with differing catchment populations, and has already been recommended by stakeholders.^31^ In some areas the size of the population fluctuates and the amount that the population travels affect the data and consideration of this requires methodological development. Normalizing by WW flow can be important in areas affected by heavy rainfall, but again the precise data requirements and methods available to account for this require development and is an ongoing area of research.

If available, clinical surveillance is considered by the interviewees to be crucial to include in dashboards, and supports conclusions made elsewhere.^31^ Clinical surveillance may include the number of cases, deaths, hospital beds, etc. However, if there is intense and frequent clinical sampling occurring in a population, WW data will show the exact same conclusions. As a result, WBE may not be an early warning system unless there is a significant uncertainty in clinical sampling or delays in accessing complete clinical data.

The inclusion of SARS-CoV-2 variant surveillance is important only if it will influence health policy changes. This is dependent on how regional health officers are planning to respond to changes in variants. The interviews expressed that while variant data is valuable, the information is perhaps more academic, although early indication of variants with additional vaccine resistance would be useful to know. The risk of misinterpreting variant analysis due to lack of understanding can be problematic, especially in public facing dashboards, and suitable guidance for data interpretation is needed.

### Implications

From the three themes analysed in the present study certain recommendations can be made to facilitate development of dashboards that are interoperable. Access to displayed data on the dashboard should not be restricted and should include sources. The preferred time-intensive graphic needs to represent a pre-determined time, focussing on recent weeks. Based on all the analysed factors that may influence geographical representation the spatial design may not be practical currently, especially if there are concerns around security and identifiable factors that are dependent on sewer-catchment locations. However, if stakeholders regard geographical comparisons as important, investments should be made to develop a way forward that considers ethical and security concerns. Clinical surveillance should be included in a dashboard, especially because WW is still classified as supplementary to this data. Similarly, there needs to be more research done with variant surveillance because there is still hesitancy around the value of providing health policy makers with this data. Standardization of dashboards may not necessarily be a research or public health priority. Instead, evaluation of approaches helps to establish best practice, which may of course result in more uniformity is dashboards, but allows for innovation and adaptation.

A strength of the qualitative approach adopted in this study is the generation of data on the processes which shaped the development of dashboards in practice. This allows us to see the emergent and iterative nature of the innovation, as well as how the data translations afforded by different dashboards are subject to multiple interpretations on account of their data and use contexts. Qualitative interviews are inevitably oriented to the generation of accounts that are situated in their specific local contexts, and thus may not have generalisability beyond these and the perspectives of the participants involved. In our study, interviews have generated findings of generic value when considering to what extent standardisation is possible and feasible when translating WW analyses into data presented via dashboards. A limitation of this study is that the initial coding of transcribed interview data informing the discussion of analytical themes were generated by one person (DM). As a rapid response study to an intervention development, the study was inevitably limited in its sample size and recruitment potential.

This study provides insight on how dashboards were developed during an acute period of a pandemic, and highlights best practice that was developed along the way. The varying experience of research groups and the initial reactions of governing bodies during the early days of the COVID-19 pandemic impacted the development of SARS-CoV-2 WW dashboards. Although in the beginning countries encountered varying degrees of challenges, today countries across Europe and North America remain focused on improving the future of WW surveillance.

## Supporting information

COREX checklist

## Data Availability

Due to the potential identifiability of participants, interview data are not available for further study. The interview questions are available in the Appendix.

## Reporting Guidelines

We adhere to the COREQ checklist for qualitative research (Appendix 3).

## Consent

Informed consent was obtained from all participants as part of study sign up.

## Author Contributions

KMO and DM conceptualised the study. DM developed the study protocol, carried out the data collection and interpreted the results. KMO supervised the study. DM wrote the first draft of the manuscript, where all authors (DM, TR and KMO) edited the manuscript and agreed on the conclusions made.

## Acknowledgements

This project was undertaken as part of MSc study at the London School of Hygiene and Tropical Medicine in 2022-23. The authors are very grateful to the 14 participants that were willing to be interviewed and take time out of their busy schedules.

## APPENDIX

### Appendix 1: Links to Dashboards used by Participants

**Table 3:** Links to Dashboards used by Interviewed Participants.

Links to the Dashboards that Participants Have the Most Experience With...
|  |
| --- |
| https://www.thl.fi/episeuranta/jatevesi/WW_weekly_report.html |
| https://searchcovid.info/dashboards/WW-surveillance/ |
| https://WWmonitoring.fuse.arup.com/ |
| https://scotland.shinyapps.io/phs-respiratory-covid-19/ |
| https://WW-observatory.jrc.ec.europa.eu/ |
| https://abwassermonitoring.at/ |
| https://www.hpsc.ie/az/respiratory/coronavirus/novelcoronavirus/surveillance/WWsurveillanceprogramme/ |
| https://coronadashboard.rijksoverheid.nl/ |
| https://covid.cdc.gov/covid-data-tracker/#WW-surveillance |
| https://pure.qub.ac.uk/en/clippings/WW-covid-work-sars-cov-2-WW-surveillance-using-gi |
| https://www.fhi.no/en/in/surveillance/overvaking-smittsomme-sykdommer-i-avlopsvann/results-from-WW-surveillance/ |

### Appendix 2: Interview Guide

Questions for Stakeholders

**WW surveillance of SARS-CoV-2**

**Environmental Surveillance & WW Dashboard Questions [prioritise the Qs with **]**

Objective 1: Development of WW Dashboards

1. ** The current Covid-19 situation has drastically changed over the last three years and thus dashboards for SARS-CoV-2 have changed from an early warning alarm system to a cost-effective system that monitors for current trends of the virus. Can you speak briefly speak about your background and your overall experience with the SARS-CoV-2 dashboards?

i. More specifically, how often they were used in your line of work?
ii. What are the strengths and weaknesses of current dashboards?
2. ** As a developer / stakeholder, please describe the process behind development of the dashboard? [*prompt qs below*]
  1. Was it an iterative process?
  2. What do you think worked well?
  3. What challenges were there that were not foreseen?

Objective 2: Standardization of WW Dashboards

1. ** A dashboard has a limited capacity to display real time information and thus sometimes only one graphic can be included. With such a limited space of information how would you hierarchise the epidemiological data to present the most valuable information displayed with the least amount of time invested by a stakeholder?

Such as a graphic demonstrating a temporal/intensity graphic of the covid-19 virus or a spatial distribution of the virus. What information should be prioritized/ guide me through what information you look for when the data are presented like this?

#### Map

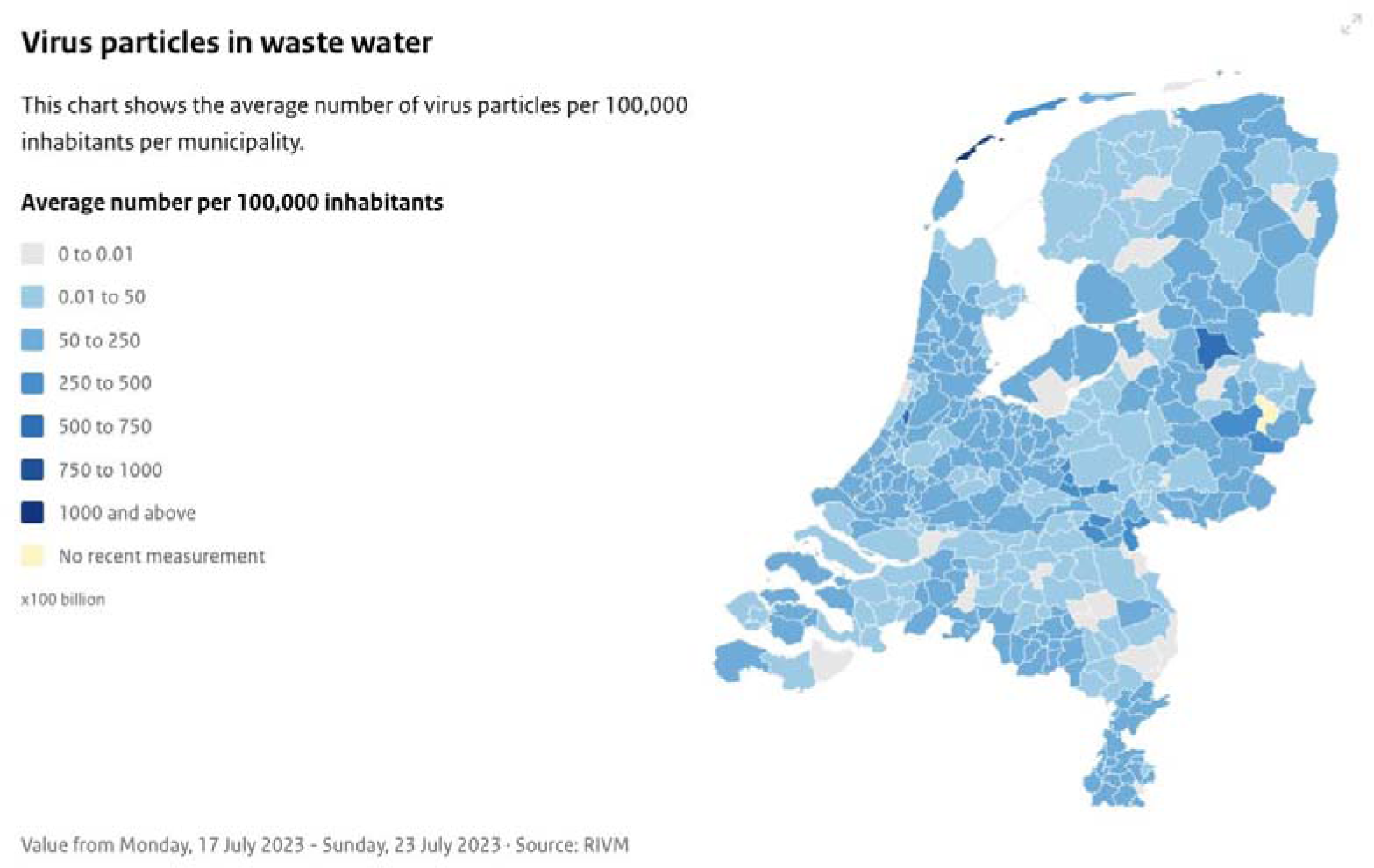

#### Intensity Time Series Example

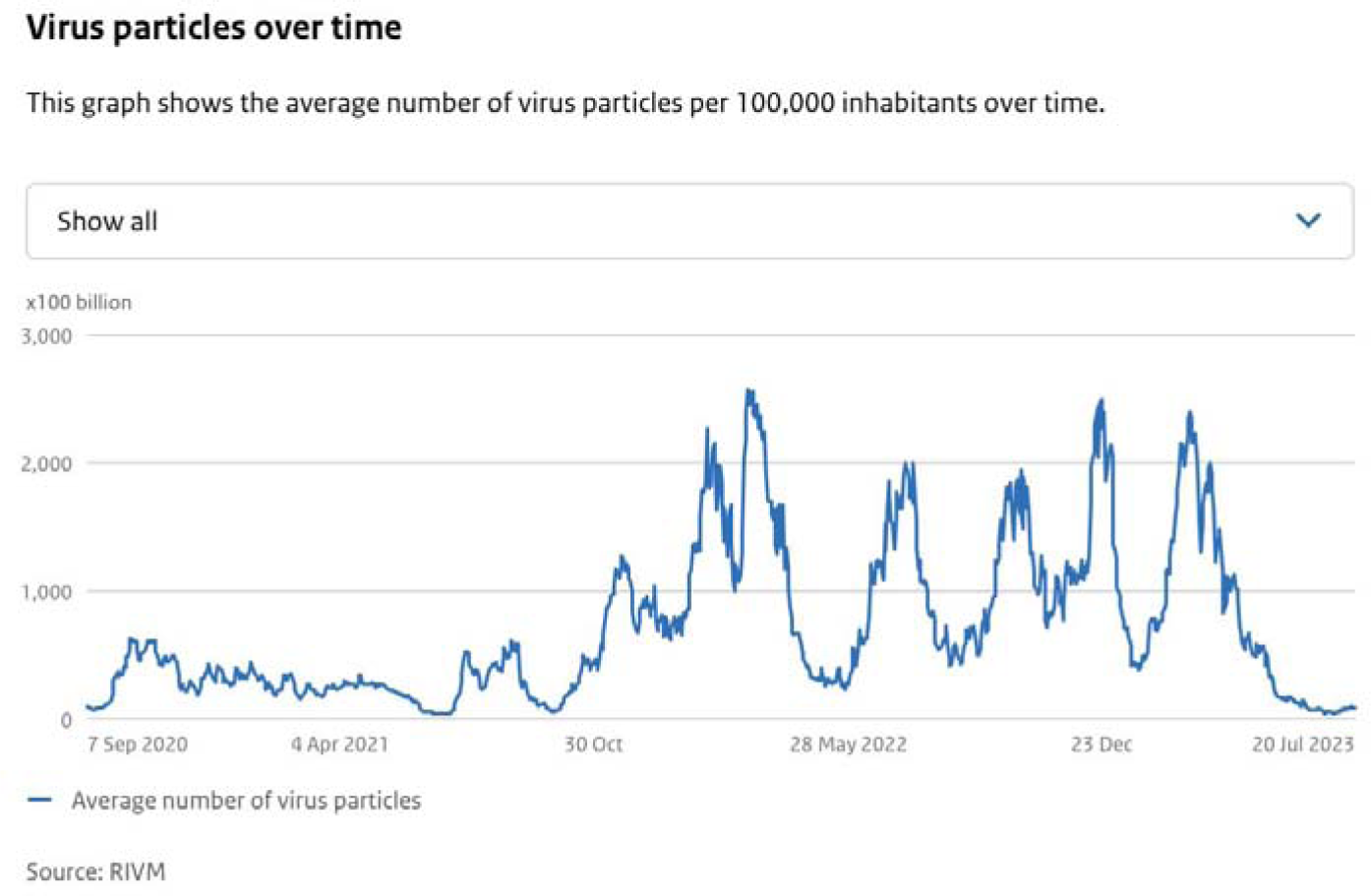

#### Questions for Stakeholders

2. 2. The information present on dashboards should ideally be standardized. When reporting the data, it was found that dashboard units can be grouped into

1. Viral Concentrations gc/L
2. Viral Concentration normalized by population size sampled.
3. Viral Concentration normalized by time
4. Viral Concentration normalized by a faecal indicator
5. A qualitative metric from the quantitative data (eg. low, medium, high)

Considering these options and the fact that they answer different epidemiological questions when accessing the dashboard, what should be the standard group and then more specifically the standard unit when representing the RNA copies of SARS-CoV-2?

Why have you chosen this option instead of the others? How would you rank the order of the units mentioned for dashboard analysis?

3. Some dashboards represent Covid-19 with a control that estimates the concentration of human faecal presence with a sample such as with the pepper mild mottle virus (PMMoV). Do you consider this an important comparison and believe that the SARS-CoV-2 should be reported in conjunction with a control virus to standardize the WW sample?

Objective 3: Gaps in Knowledge of what needs to be included within WW Dashboards

##### Epidemiological/Clinical Surveillance

1. ** Should a dashboard include the activity of the virus in the community in question including positive cases, hospitalizations, deaths, etc.

1. Is this currently carried out in your country?
2. If you think it should be done, what prevents this?

##### Variant Surveillance

2. Should variant be included in a dashboard?

1. How is this information used by you?
2. Why do you find it useful (or not)?

##### Data Transparency

3. As a stakeholder how crucial is it to have downloadable source data on a dashboard? Should this data be available for others?

a. If this is deemed as necessary, what kind of specific information should be included in the downloads: the population, the number of samples taken etc.?
4. ** Many dashboards accessed during this research are public domain, and with this in mind do you consider that dashboards should include guidance for interpretation and analysis of the data? Should access to the dashboards be limited to public health officials and experts and offer restricted access?

On a scale from 0 - 5 should the information be geared towards the public domain (0) or public health experts (5)
1- Feasible access to the Public
2- Local Community leaders and Media
3- Community Health Workers and Primary Care Providers
4- Healthcare Providers in Leadership Positions for Health Systems 5-Stake holders making decisions or allocating resources
a. Why have you selected your answer?

Final Q:** **Is there anything else that you wish to raise?**

